# Systematic identification of rare disease patients in electronic health records enables evaluation of clinical outcomes

**DOI:** 10.1101/2025.05.02.25325348

**Authors:** Arjun S. Yadaw, Eric Sid, Hythem Sidky, Chenjie Zeng, Qian Zhu, Ewy A. Mathé, the N3C Consortium

## Abstract

**Background:** Identifying rare disease (RD) patients in electronic health records (EHR) is challenging, as more than 10,000 rare diseases are not typically captured by clinical coding systems. This limits the assessment of clinical outcomes for RD patients. This study introduces a semiautomated approach to map RDs to appropriate codes, that is applicable across various EHR systems. By improving RD patient identification, this method facilitates the analysis of clinical outcomes and disease severity in the RD population. We exemplify this by utilizing large EHR datasets such as those in the National COVID Cohort Collaborative (N3C) with over 21 million patients.

**Methods:** We developed a semiautomated workflow to enumerate RD-specific SNOMED-CT and ICD-10 codes, starting with 12,003 GARD IDs mapped to ORPHANET. This process linked RDs to SNOMED-CT and ICD-10 codes, applying exclusion criteria based on group of disorders. We created an extensive list of SNOMED-CT codes with descendants from the OHDSI atlas and performed phenotype filtering, removing irrelevant codes. The final list included 12,081 SNOMED-CT codes and 357 ICD-10 codes for further analysis, enabling the identification and mapping of rare diseases in EHR.

**Results:** Our semiautomated workflow identified 357 RD-specific ICD-10 codes and 12,081 SNOMED-CT codes representing 6,342 RDs which are categorized into 30 Orphanet linearization classes. We exemplify the utility of these codes by performing a preliminary univariate analysis of COVID-19 outcomes in a large cohort of 4,835,718 COVID-19 positive individuals in N3C, of which 404,735 (8.37%) were identified as having preexisting RD. The mortality and hospitalization risk ratios for rare RD classes ranged from 0.23 - 5.28 and 0.93 - 3.13, respectively (p-values <0.001).

**Conclusions:** Our systematic and automated workflow enables rapid identification of rare disease patients across diverse EHR systems. We demonstrate its utility by evaluating COVID-19 severity outcomes by rare disease classes in the N3C cohort. These findings support the need for targeted preventive healthcare interventions and highlight the potential for future research on long COVID, COVID-19 reinfection, and other outcomes in the rare disease population.

## Introduction

Currently, 25 to 30 million individuals in the United States (US) and over 300 million people across the world are diagnosed with a rare disease (RD)^1–3^. This means that 3.5% to 5.9% of the world’s population is affected by RDs^3^. Collectively, rare diseases represent a significant public health burden with more than 10,000 identified RDs^1^. Among them, only 5% have FDA approved therapeutic options^1^. Indeed, developing interventions for a single RD is very costly and timely, and drug development costs are recouped by a limited number of patients, resulting in markedly higher costs of orphan drug treatments than non-orphan drugs, particularly in the US^4^. Notably, our ability to identify and evaluate this patient population as a whole or as groups, rather than one disease at a time, provides opportunities for finding shared interventions. It thus becomes possible to leverage shared information about diseases, including similarities in genetic or other molecular profiling, mechanisms or action, symptoms, to group them for drug repurposing efforts or implementation of basket trials^5^. In this work, we focus on identifying RD patients in electronic health records (EHRs), as these provide a rich source of information for translational research efforts that aim to understand disease patterns, find new potential treatments and the guide implementation of clinical trials^6–8^.

The identification of RD patients in EHRs remains challenging due to the heterogeneity of the coding systems used across hospital systems and the lack of collection of RD based codes^9^. In the US, most hospital systems rely on the Health Insurance Portability and Accountability Act (HIPAA)-compliant standardized ICD10-CM codes to report disease diagnoses^10^. Importantly, these systems are primarily put in place for billing and business assessments^11^. An independent and complementary standard nomenclature established in 1999, SNOMED-CT, provides a means for comparing clinical healthcare information across different institutions and can be mapped to the ICD10-CM^12^. In the United States, the ICD10-CM and SNOMED codes are heavily used in EHR systems to capture diagnosis-related information and standardized complementary information on diseases, respectively. Another highly utilized and relevant standard is the Observational Medical Outcomes Partnership (OMOP) Common Data Model (CDM)^13^ which produces standardized vocabularies that group differing medical terms across disparate clinical systems into a standardized concept. Using such standardized concepts enables the integration and cross-evaluation of data across different systems, each of which collects data via its own standards and criteria.

In parallel, large national and international initiatives are providing detailed, complementary information on RDs to further enhance data cross-linking and definitions of diseases. In the United States, the GARD (Genetic and Rare Diseases Information Center) provides access to up-to-date research, resources and diagnostic guidance to RD patients^1^. The Monarch Initiative^14^, an international consortium aggregating and harmonizing resources on human genes and diseases, model organisms, genomic data, and expression/pathway information, produces MONDO (Monarch Merged Disease Ontology)^15^. MONDO provides an ontology for connecting diseases across databases and resources. Orphanet^16^, an international effort supported by the European Commission, provides information on RDs in many different languages as well as orphan drug and RD inventories. Importantly, these resources cross-reference each other to promote interoperability and cross-sharing of information, which enables the RD code generation introduced in this study^1,12,16,17^.

There are two broad approaches to identifying RD patients in EHRs, those based on knowledge mining of EHR records and those based on the use of standardized codes. One example of the former includes the use of EHR records to learn text embeddings, which are in turn used to predict the likelihood of patients having an RD or lipodystrophy in this case^18^. In another study, acute hepatic porphyria patients were predicted via support vector machines with information on diagnosis, medications, procedures and clinical notes as input^19^. AI-based digital health assistants have also been applied to identify RDs in EHR records, largely using symptom data^20,21^. In contrast, other studies rely heavily on standardized codes. For example, one study performed on Asian healthcare system data utilized a value set of SNOMED codes with demographic and symptom filters to evaluate patients with Fabry disease and familial hypercholesterolemia (rare genetic disease)^22^. Another recent study developed algorithms to identify patients with Goucher disease (GD) in an EHR system using ICD codes and standardized laboratory results and symptoms^23^. Globally, these studies aim to ameliorate the process of identifying RD patients in EHR records to help shorten the diagnostic odyssey length.

Nonetheless, it is worth noting that a recent study comparing the performance of LLMs in diagnosing diseases demonstrated that more traditional methods still outperforming LLMs^24^. Furthermore, predictions via knowledge mining are typically focused on single diseases and it is currently not feasible to develop such an algorithm for all RDs. Furthermore, while the use of standardized codes is applicable to a broader set of diseases, a comprehensive starting list of disease codes is needed. We note that there is currently no gold standard covering more than 10K existing RDs that are EHR system types agnostic. To the best of our knowledge, no algorithm to date has aimed to identify all US-defined RDs in EHRs.

In this study, we aimed to develop an automated process for generating RD-specific ICD-10 and SNOMED-CT codes for systematic identification of RD patients in EHRs. We recognize the continued impact of COVID-19 with more than 7.09 million deaths worldwide, as of February 23^rd^, 2025^25^ and thus exemplify the utility of this list of RD-specific codes to assess the potential impact of COVID-19 on RD patients. We mapped these RD-specific codes to OMOP concepts to identify RD patients in the National COVID Cohort Collaborative (N3C)^26,27^. Patients with RD were further classified into Orphanet linearization groups to evaluate their univariate relative risk of COVID-19 severity outcomes. Preliminary findings from our analysis demonstrate the utility of this mapping for evaluating clinical outcomes within the rare disease population using EHR records, particularly in the context of the COVID-19 outcomes. By establishing a standardized and comprehensive framework for rare disease identification, this work lays the groundwork for a wide range of downstream analyses, including assessments of long COVID, COVID-19 reinfection, antiviral treatment effects, and other pandemic-related outcomes in vulnerable rare disease cohorts.

## Methods and Materials

### Development of the RD phenotyping algorithm

As a starting point, we compiled a curated RD list of 12,003 comprising GARD IDs, where each represents a unique RD. The GARD maintains a comprehensive registry of RDs, with particular emphasis on conditions meeting the United States regulatory definitions of rare disease status. This registry incorporates external citations to authoritative sources regarding disease etiology, diagnostic criteria, and therapeutic management protocols.^1^These GARD IDs were mapped to Orphanet concepts (July 2023 version)^16^ which have been mapped to SNOMED-CT and the ICD-10.

These retrieved SNOMED-CT and ICD-10 mappings then underwent a series of filtration steps to produce the final list of RD-specific codes. Prior to converging to a final RD phenotypic algorithm, four semiautomated approaches were evaluated prior to converging to the final algorithm. These iterations are described in the Supplementary Materials and in (**Supplementary Figure 1**).

In the final algorithm, we first removed RDs associated with the tag of “group of disorders” (e.g., Disorder of carbohydrate metabolism has 396 descendent record counts and is thus tagged as “group of disorders”) on the basis of the Orphanet definition. Second, we expanded the list of codes to include descendent concepts of our remaining 6,327 SNOMED-CT codes by searching for SNOMED-CT codes using the Observational Health Data Sciences and Informatics (OHDSI) Atlas tool^28^. We assumed that parent concepts are RDs and that their corresponding descendant concepts are thus also RDs. We then curated descendent concepts to filter out those concepts that were represented in the Human Phenotype Ontology (HPO)^29^ via the HPO API. Filtering out phenotypes ensures that the resulting codes represent the specific disease itself rather than manifestations or characteristics of the disease which could be shared across multiple diseases (e.g., sensory motor neuropathy, muscle weakness, etc.). The application of these final filtering steps resulted in the final list of RD-specific codes that can be utilized to identify RDs in any EHR system. See further details in the Supplementary Materials.

The resulting SNOMED-CT codes from all 4 phenotypic algorithms were evaluated against a manually curated list. To create this list, we conducted comprehensive manual reviews by cross-referencing multiple authoritative rare disease knowledge bases to verify disease classification as rare. In addition to the estimated prevalence in the US general population, the prevalence of each disease in large scale population-based cohorts was also considered during this verification process.

### Characterization of RDs

The Orphanet linearization classification, accessed through Orphadata (July 2023 version), was used to produce groups of RDs with similar etiologies. The RD-specific SNOMED-CT and ICD-10 codes were directly mapped to the Orphanet linearization. When a disease is present in multiple classes, the linearization rules for Orphanet classifications prioritize the most severely affected body system, the most determining involvement for prognosis, and finally the specialist most likely to be relied on for the management of the disease^30^. The number of RDs mapped to the linearization is based on the number of unique GARD IDs prior to phenotype filtering.

We also retrieved RD related publications from the Rare Disease Alert System (RDAS) [https://rdas.ncats.nih.gov], an RD based integrative research data platform, to access the research effort made on those RDs. GARD IDs were applied to search for relevant publications via the RDAS API [https://rdas.ncats.nih.gov/apis/publications].

### Establishing the COVID-19 cohort in the N3C enclave

The National COVID Cohort Collaborative (N3C)^15^ is an aggregated clinical data resource with participating data partners in the U.S., harmonized using the Observational Medical Outcomes Partnership (OMOP) data model, and subjected to quality reviews and checks. All patients within the N3C Enclave possess historical data from the same healthcare system dating back to January 1, 2018. This dataset includes information on preexisting health conditions (e.g., comorbidities) and other relevant medical history (lookback data)^31^. For this study, we used N3C enclave data tables from version V154 which includes 21,704,702 patients. Patients with COVID-19 were identified as those with a positive COVID-19 diagnosis, based on reverse transcription polymerase chain reaction (RT-PCR) or Antigen (Ag) or U07.1 diagnosis tests, between 1^st^ January 2020 and 4^th^ January 2024. The following exclusion criteria were then applied to construct the COVID-19 cohort in this study: 1) patients with missing or invalid data on sex (missing) and age (missing or ages ≤ *1*) were removed; 2) patients with no encounter visit before and after COVID-19 diagnosis date were removed; and 3) removed records from data partners with data that did not meet N3C quality control criteria (**Supplementary Figure 2**). The N3C Data Enclave is approved under the authority of the National Institutes of Health Institutional Review Board. Each N3C site maintains an institutional review board–approved data transfer agreement.

### Identifying RD patients in the COVID-19 cohort

We map our RD-specific SNOMED-CT codes to the OMOP concept table to obtain the corresponding OMOP concept IDs. These were then linked, along with the ICD-10 codes, to the condition occurrence table, allowing us to identify patients diagnosed with a rare disease (RD) prior to their COVID-19 diagnosis within our COVID-19 cohort. An additional RD incidence filter was applied to remove diseases with an incidence rate greater than 6/10,000, following the definition of RD in the United States^1^. Furthermore, patients associated with the Orphanet linearization class of “rare disease due to toxic effects” were removed, because this linearization contained a mix of etiologies and disease types (e.g., complications from a medical product, environmental exposures, substance use and abuse), rendering interpretation of this group difficult. Patients associated with the “rare transplantation disease” group were removed because the group was limited to diseases that are a comorbidity or complication of having had a transplant (e.g., aneurysm of the vein of the transplanted kidney). Other patients with “rare odontologic disease” and “rare abdominal surgical disease” were removed due to missing mortality information for those patients available in N3C. The “Rare genetic disease” and “rare infertility” classes were not evaluated because no patients mapped to those classes.

### COVID-19 Outcomes and demographics

Unadjusted mortality and hospitalization were evaluated as binary variables. Mortality was defined by intersecting patients who were reported as dead (any cause) with patients who were deceased upon discharge from their COVID-19 visit. Hospitalization was defined as positive for patients with a hospitalization stay within 16 days of the date of COVID-19 diagnosis ^13,16^.

The demographic variables evaluated included: 1) age in years categorized as 1-20, 21-40, 41-65, >65years and 2) body mass index (BMI, in *kg*/*m*^2^ ) categorized as underweight (<18.5), normal (18.5 - 25), overweight (25 - 30), BMI obese (≥30); 3) sex (Male, Female); 4) self-reported race (Asian, Black or African American, White, missing/unknown); 5) ethnicity (Hispanic or Latino, Non Hispanic or Latino, missing/unknown); 5) smoking status (Current or former smoker, Non-smoker). The baseline characteristic demographic table (Table 1) was created by using the R libraries (gtsummary (2.1.0), tidyverse (2.0.0) & gt (0.11.1) libraries)^32^

**Table 1:**
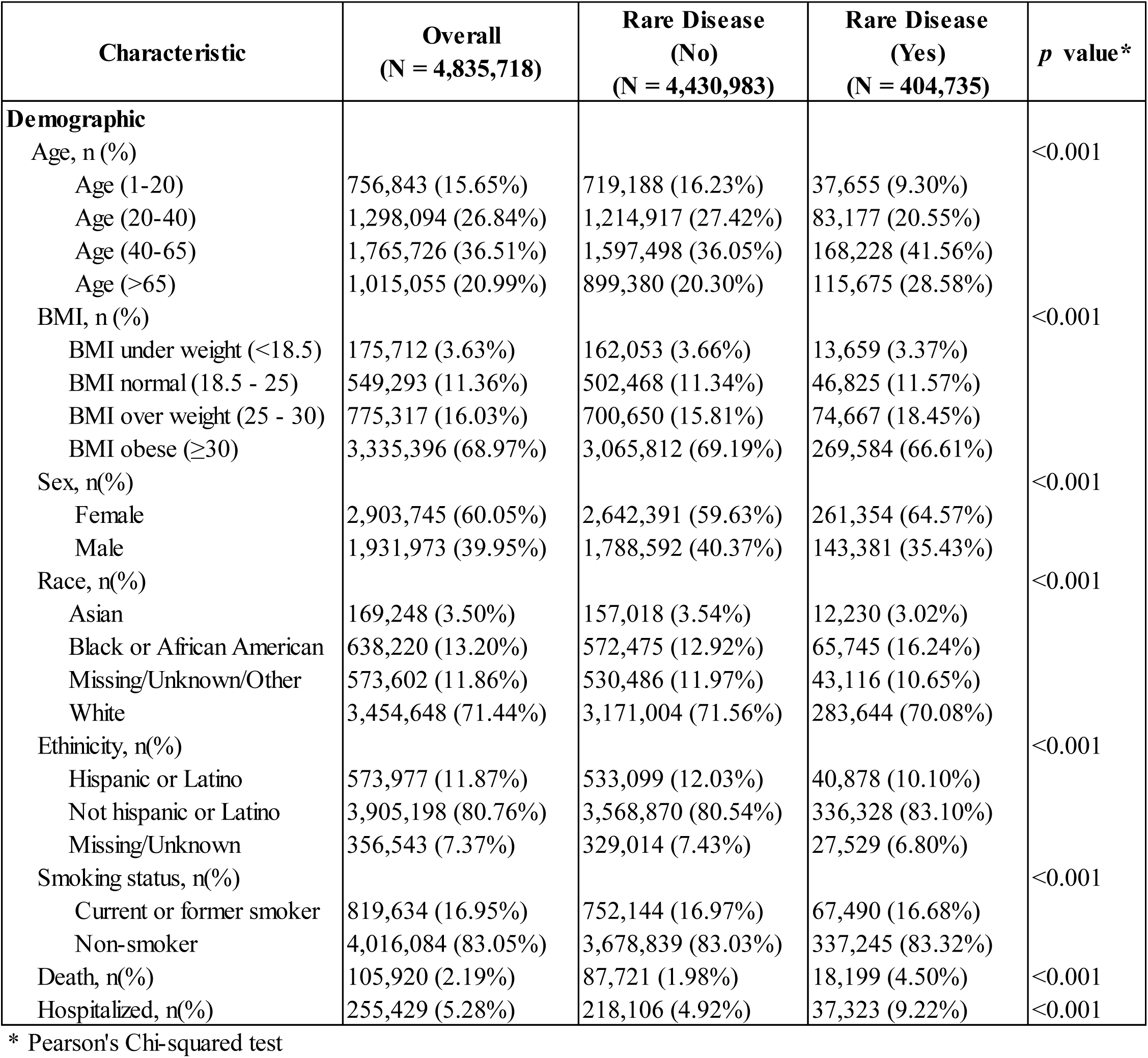
Characteristics of COVID-19 patients identified within the N3C enclave stratified by those with or without preexisting RD. Distribution of all covariables differ between patients with/without RD Chi-square test for categorical variables p-values <0.001).

| Characteristic | Overall<br>(N = 4,835,718) | Rare Disease<br>(No)<br>(N = 4,430,983) | Rare Disease<br>(Yes)<br>(N = 404,735) | *p* value* |
| --- | --- | --- | --- | --- |
| **Demographic** |  |  |  |  |
| Age, n (%) |  |  |  | <0.001 |
| Age (1-20) | 756,843 (15.65%) | 719,188 (16.23%) | 37,655 (9.30%) |  |
| Age (20-40) | 1,298,094 (26.84%) | 1,214,917 (27.42%) | 83,177 (20.55%) |  |
| Age (40-65) | 1,765,726 (36.51%) | 1,597,498 (36.05%) | 168,228 (41.56%) |  |
| Age (>65) | 1,015,055 (20.99%) | 899,380 (20.30%) | 115,675 (28.58%) |  |
| BMI, n (%) |  |  |  | <0.001 |
| BMI under weight (<18.5) | 175,712 (3.63%) | 162,053 (3.66%) | 13,659 (3.37%) |  |
| BMI normal (18.5 - 25) | 549,293 (11.36%) | 502,468 (11.34%) | 46,825 (11.57%) |  |
| BMI over weight (25 - 30) | 775,317 (16.03%) | 700,650 (15.81%) | 74,667 (18.45%) |  |
| BMI obese (≥30) | 3,335,396 (68.97%) | 3,065,812 (69.19%) | 269,584 (66.61%) |  |
| Sex, n(%) |  |  |  | <0.001 |
| Female | 2,903,745 (60.05%) | 2,642,391 (59.63%) | 261,354 (64.57%) |  |
| Male | 1,931,973 (39.95%) | 1,788,592 (40.37%) | 143,381 (35.43%) |  |
| Race, n(%) |  |  |  | <0.001 |
| Asian | 169,248 (3.50%) | 157,018 (3.54%) | 12,230 (3.02%) |  |
| Black or African American | 638,220 (13.20%) | 572,475 (12.92%) | 65,745 (16.24%) |  |
| Missing/Unknown/Other | 573,602 (11.86%) | 530,486 (11.97%) | 43,116 (10.65%) |  |
| White | 3,454,648 (71.44%) | 3,171,004 (71.56%) | 283,644 (70.08%) |  |
| Ethnicity, n(%) |  |  |  | <0.001 |
| Hispanic or Latino | 573,977 (11.87%) | 533,099 (12.03%) | 40,878 (10.10%) |  |
| Not hispanic or Latino | 3,905,198 (80.76%) | 3,568,870 (80.54%) | 336,328 (83.10%) |  |
| Missing/Unknown | 356,543 (7.37%) | 329,014 (7.43%) | 27,529 (6.80%) |  |
| Smoking status, n(%) |  |  |  | <0.001 |
| Current or former smoker | 819,634 (16.95%) | 752,144 (16.97%) | 67,490 (16.68%) |  |
| Non-smoker | 4,016,084 (83.05%) | 3,678,839 (83.03%) | 337,245 (83.32%) |  |
| Death, n(%) | 105,920 (2.19%) | 87,721 (1.98%) | 18,199 (4.50%) | <0.001 |
| Hospitalized, n(%) | 255,429 (5.28%) | 218,106 (4.92%) | 37,323 (9.22%) | <0.001 |

\* Pearson's Chi-squared test

### Statistical assessments

Mortality and hospitalization risk ratios (RRs) were calculated via the following formula:

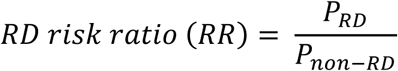

where *P*_*RD*_ and *P*_*non*−*RD*_ are the probabilities of the event (mortality/hospitalization) occurring in preexisting RD patients and non-preexisting RD patients respectively. RR = 1: The risk of the event is the same in both groups (no effect), RR > 1: The event is more likely in the exposed group (increased risk). RR < 1: The event is less likely in the exposed group (decreased risk or protective effect). Statistical significance was defined for p-values < 0.05 and 95% confidence intervals around the estimates of RRs. Statistical modeling was conducted within the N3C enclave using SQL, Python (3.10.16), statsmodels (0.14.4), Patsy (1.0.1), Numpy (1.26.4), Pandas (1.5.3) and Spark SQL (3.4.1.34).

## Results

Our main goal is to build a semiautomated phenotyping algorithm that produces RD-specific ICD-10 and SNOMED-CT codes (**Figure 1**). These codes are key to identifying RD patients in any EHR system that leverages them, thereby enabling analyses of numerous RDs together. We exemplify this utility in a COVID-19 case study where we conducted a clinical assessment of RD patients in the N3C Enclave, including COVID-19 related mortality and hospitalization (**Figure 1**).

**Figure 1:**
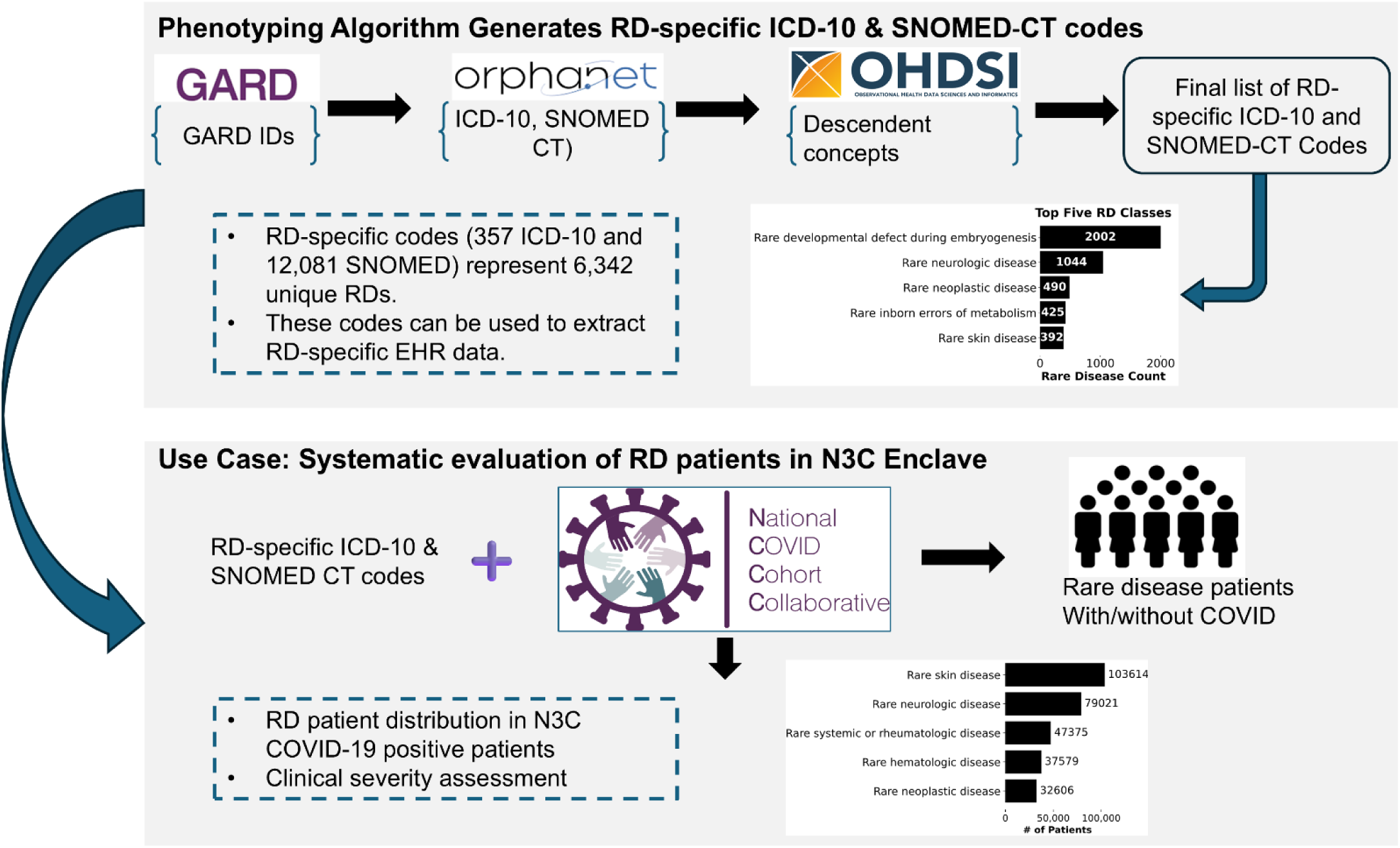
Study design. In this study, we developed a phenotyping algorithm that leverages GARD, Orphanet, and OHDSI to produce RD-specific ICD-10 and SNOMED-CT codes. These codes are specific to individual diseases are devoid of group disorders and phenotypes. We demonstrate the utility of these codes through a clinical assessment of RD patients in the N3C Enclave. Specifically, we systematically identified RD patients in the enclave and evaluated distributions of RD patients by their Orphanet class, as well as risk of COVID-19 mortality.

### Phenotyping algorithm for the systematic identification of RD patients in EHR systems

As a starting point, a list of RDs, based on the US definition, was derived by mapping a curated list of 12,003 GARD IDs to Orphanet, resulting in 9,369 RDs with 6,553 SNOMED-CT codes and 575 ICD-10 codes. Four algorithms were subsequently tested (**Supplementary Figure 1**), and for each, various combinations of filters were applied to remove phenotypes, groups of diseases and common diseases. Our goal was to produce a final algorithm that reduced the reliance on manual curation while retaining as many RDs as possible. An evaluation of the commonalities in the resulting ICD-10 and SNOMED-CT codes between the four algorithms revealed that the codes dropped in the final algorithm largely represented groups of disorders or phenotypes (**Supplementary Figure 3**). We further compared the resulting SNOMED-CT codes from our semiautomated approaches to those obtained from a manually curated list of 1,715 diseases (**Supplementary Table 1**). Importantly, our final approach showed the largest percentage of diseases mappable in our manually curated list at 89.8%. Of these,11.6% were determined not RD (false positives), leaving the large majority, 88.4% as true RDs. While other approaches greatly minimize this false positive rate (as low as 1.0%), this approach comes at the cost of low mappability (a low as 42.6%). We conclude that overall, our final algorithm (**Figure 2**) minimized the amount of manual curation, number of phenotypes and number of groups of diseases, and number of false positives resulting in an optimized approach to producing RD-specific codes for the identification of RD in EHRs. Of the initial 6,553 SNOMED-CT and 575 ICD-10 codes, 112 ICD-10 and 226 SNOMED-CT codes were removed because they are associated with a group of disorders, and 106 ICD-10 and 488 SNOMED-CT codes were removed because they are associated with phenotypes (as per available HPO mapping) rather than diseases. In total, a final list of RD-specific codes comprised 357 ICD-10 codes and 12,081 SNOMED-CT IDs, representing a total of 6,342 unique RDs. This list is available in (**Supplementary Table 2)**, along with associated GARD IDs, names and Orphanet linearization classes and further details on each filtering step are found in the **Supplementary Materials**.

**Figure 2:**
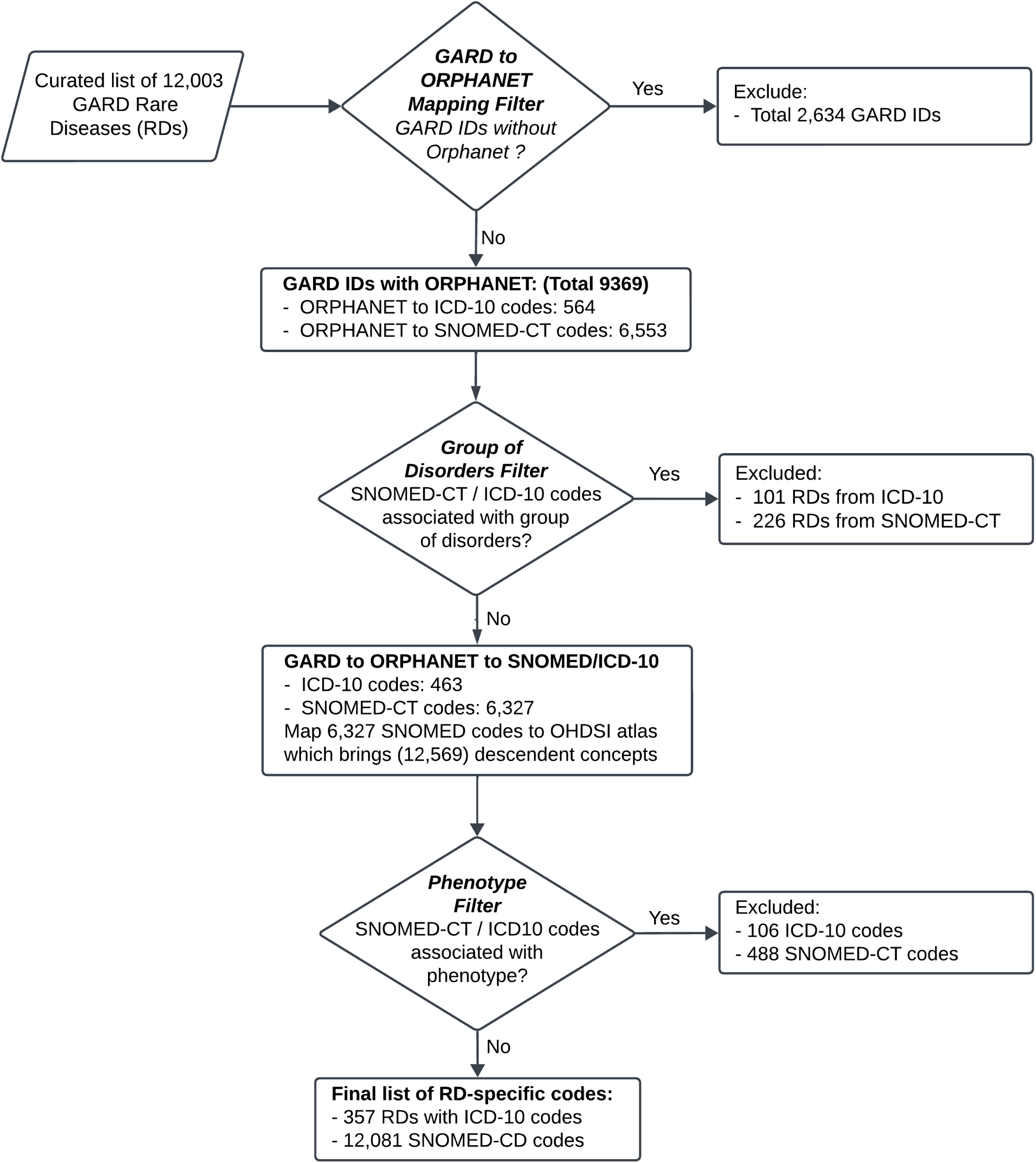
RD phenotyping Algorithm. A semi-automated algorithm was built to define RD-specific ICD-10 and SNOMED-CT codes. The algorithm starts with 12,003 RDs, each defined by a unique GARD ID, which are mapped to Orphanet to retrieve associated ICD-10 and SNOMED-CT codes. At this step, groups of disorders and broad concepts (e.g., one-to-many mappings) are removed. Next, a phenotype exclusion step is performed after pulling in descendent concepts of the remaining SNOMED-CT codes. As a result, 357 ICD-10 codes and 12,081 SNOMED-CT codes are produced, representing at least 6,342 RDs.

### Characteristics of disease represented by RD-specific SNOMED/ICD-10

At least 6,342 diseases (estimated prior to the phenotype filter) with RD-specific codes were categorized via Orphanet linearization (**Supplementary Table 3**). Because each Orphanet RD maps to a single Orphanet linearization, these categories are useful for evaluating groups of diseases and for clinical assessment. (**Figure 3A)** depicts the number of RDs per category for the 30 linearizations that represent the 6,342 diseases. Several linearization classes are poorly represented with < 10 codes. These underrepresented linearizations include rare urogenital, rare maxilo-facial surgical, rare abdominal surgical, rare infertility, rare genetic, rare surgical thoracic, rare transplantation, and rare surgical cardiac diseases.

**Figure 3:**
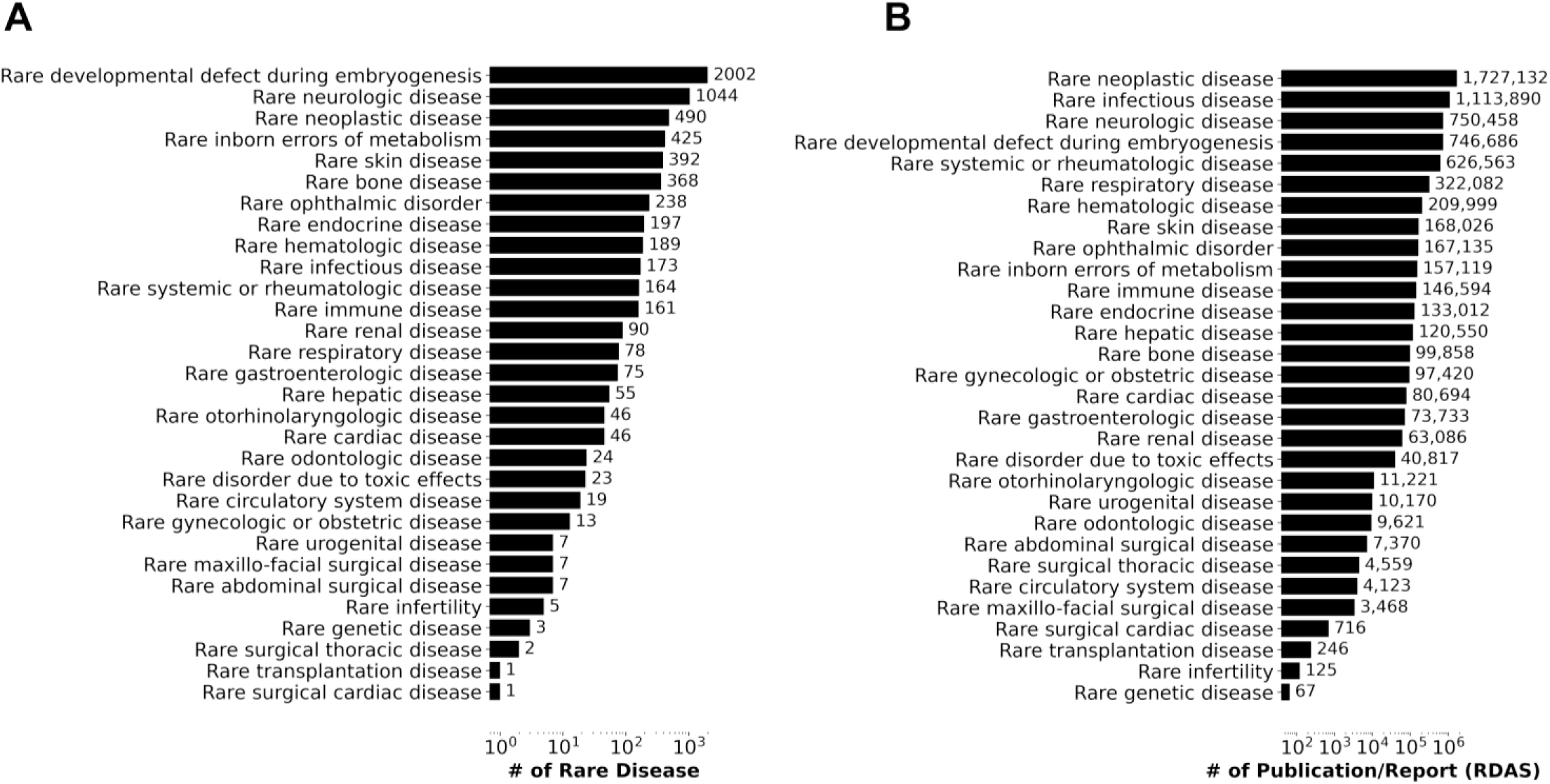
Characteristics of the 6,342 unique RDs represented by RD-specific ICD-10 and SNOMED-CT codes. A) Number of diseases by Orphanet linearization; B) Number of publications/reports from (RDAS).

The most well represented classes of RD are “rare developmental defects during embryogenesis” and “rare neurologic disease" with 2002 and 1,044 RDs in each, respectively. The least represented linearizations include rare urogenital, rare maxillo-facial surgical and rare surgical thoracic diseases, with fewer than 10 ICD-10 and SNOMED-CT codes. The remaining linearization classes include 13 to 490 unique codes.

Interestingly, the number of representative codes for each linearization aligns well with how well these diseases are evaluated in the literature. (**Figure 3B)** shows the number of associated reports/publications obtained via Orphanet linearization. The most widely reported (with > 750,000 publications/reports) categories include rare neoplastic, rare infectious and rare neurological diseases, which are also the top 3 represented linearizations by our resulting RD-specific codes. The 3 least represented linearizations in the literature, with fewer than 250 publications are rare transplantation, infertility and rare genetic disease.

### Use case: evaluation of RD patients in a COVID-19 cohort using the N3C enclave

We exemplify the utility of our list of RD-specific codes by studying a large number of patients with RDs in the N3C enclave which comprises 21,704,702 patients, with 8,463,370 having a positive COVID-19 diagnosis (**Supplementary Figure 2, Methods**). After applying exclusion criteria based on missing data on age and sex, <1 encounter visits before/after COVID-19 diagnosis date, and data that did not pass N3C quality control, our final COVID-19 cohort comprised 4,835,718 patients. As a final step, we further applied filtering criteria based on incidence, ensuring that only diseases with an incidence rate less than 6/10,000 were represented. In total, 7 ICD-10 codes and 18 SNOMED-CT codes were removed (**Supplementary Figure 4**).

We next cross-linked our list of RD-specific codes resulting from our phenotypic algorithm to stratify patients into those with or without a preexisting RD diagnosis. Of the 12,081 RD-specific codes identified by our algorithm, 10,611 were mappable to N3C, including 350 ICD-10 codes and 12,063 SNOMED-CT codes. In total, we identified 404,735 patients (8.37%) with a preexisting RD, while 4,430,983 did not have a preexisting RD (**Supplementary Figures 2 and 4, Table 1**). Given the heterogeneity of RDs, we further stratified patients into the 23 Orphanet linearization (**Supplementary Figure 5A)**. The rare skin disease group shows the largest representation in N3C (N=103,614), followed by rare neoplastic disease (N= 79,021) and rare systemic or rheumatologic disease (N=47,375). Conversely, rare surgical disease (N= 101), the rare maxillo facial surgical diseases (N= 119), and rare circulatory system diseases (N= 727) were the least represented. The number of patients in the remaining groups ranged from 3,226 to 37,579. Notably, no patients mapped to the Orphanet linearization classes of rare genetic disease and rare infertility.

Globally, among the 404,735 patients with preexisting RD, 37,323 (9.22%) were hospitalized and 18,199 (4.50%) died. These patients with preexisting RD were more likely to be female than male (64.57% female in the RD group vs 59.63% female in the non-RD group), and their age distributions differed (for example, 9.30% under 20 years in the RD group vs 16.23% under 20 years in the non-RD group) from those of patients without preexisting RD. The demographic data are shown in Table 1, which reveals significant differences in age, BMI, race, ethnicity, and smoking status between patients with and without preexisting RD. Furthermore, higher rates of mortality (4.50% vs 1.98% in non-preexisting RD patients) and hospitalization (9.22% vs 4.92% in non-preexisting RD patients) were observed in patients with preexisting RD (**Table1**). The distribution of mortality for patients with and without preexisting RDs was also evaluated by age and sex categories to reveal potential differences between those two demographics (**Supplementary Figure 6**).

We next conducted univariate analyses to evaluate the impact of age and sex on mortality of patients with and without preexisting RD. For all age groups, we observed a higher mortality in both males and females although the increasing rate of mortality is more pronounced in males than in females (**Supplementary Figure 6**). For example, in the 65+ age group, the percent mortality difference between patients with and without preexisting RD is 4.22% higher in males, whereas it was 2.2% (Chi-square p-value < 0.0001). We also observe an increase in the percent mortality difference as patients get older. For example, male patients aged 1-20 years had a 1.14% mortality, whereas those aged 65+ years had an 11.83% mortality. Similar observations are made when evaluating the hospitalization rate.

The rates of mortality and hospitalization were then explored for each linearization class except for those classes excluded because of missing mortality heterogeneity of RD represented, or a lack of patient mapping (see **Methods**). Our analyses revealed the highest percentages of mortality and hospitalization for patients with rare neoplastic disease (10.4% mortality and 12.8% hospitalization), rare respiratory disease (9.0% mortality and 15.0% hospitalization), and rare gastroenterological disease (7.1% death and 12.5% hospitalization) **(Supplementary Figures 5B and 5C)**. The least affected linearization classes include rare skin diseases (1.6% mortality and 4.6% hospitalization), rare gynecologic or obstetric diseases (0.5% mortality and 6.2% hospitalization) and rare urogenital diseases (1.8% mortality and 5.2% hospitalization).

Finally, we aimed to evaluate the unadjusted risk of COVID-19 related hospitalization and mortality in COVID-19 patients with and without preexisting RDs (**Table 2).** The highest mortality RRs were observed for the rare neoplastic disease group (RR=5.275; 95% CI= 5.272-5.278; *p* < 0.001), the rare respiratory disease group (RR = 4.56; 95% CI= 4.556-4.562; *p* < 0.001), and the rare gastroenterologic disease (RR = 3.563; 95% CI= 3.553-3.575; *p* < 0.001) (**Table 2**). For hospitalized individuals, the three highest hospitalization RRs were observed for rare endocrine disease (RR = 3.128; 95% CI= 3.118-3.135; *p* < 0.001), rare respiratory disease (RR= 3.038; 95% CI= 3.036-3.039; *p* < 0.001), and rare hematology disease (RR= 2.885; 95% CI=2.885-2.887 *p* < 0.001) (**Table 2**).

**Table 2:** Relative Risk Ratio of Rare Disease Classes with Respect to Control (COVID-19 positive) for Severity Outcomes (Mortality/Hospitalization)

| Variables | Total Patients | Number of mortality | Mortality |  | Variables | Total Patients | Number of hospitalization | Hospitalization |  |
| --- | --- | --- | --- | --- | --- | --- | --- | --- | --- |
|  |  |  | Mortality risk ratio (95% CI) | p value |  |  |  | Hospitalization risk ratio (95% CI) | p value |
| Rare neoplastic disease | 32606 | 3405 | 5.275 (5.272 - 5.278) | <2.2E-308 | Rare endocrine disease | 3994 | 615 | 3.128 (3.118 - 3.135) | <2.2E-308 |
| Rare respiratory disease | 31549 | 2848 | 4.56 (4.556 - 4.562) | <2.2E-308 | Rare respiratory disease | 31549 | 4718 | 3.038 (3.036 - 3.039) | <2.2E-308 |
| Rare gastroenterologic disease | 8493 | 599 | 3.563 (3.553 - 3.575) | <2.2E-308 | Rare hematologic disease | 37579 | 5337 | 2.885 (2.885 - 2.887) | <2.2E-308 |
| Rare hepatic disease | 8375 | 572 | 3.45 (3.438 - 3.46) | <2.2E-308 | Rare circulatory system disease | 727 | 98 | 2.739 (2.692 - 2.789) | <2.2E-308 |
| Rare endocrine disease | 3994 | 268 | 3.389 (3.367 - 3.414) | <2.2E-308 | Rare neoplastic disease | 32606 | 4175 | 2.601 (2.6 - 2.602) | <2.2E-308 |
| Rare bone diseases | 4113 | 255 | 3.132 (3.11 - 3.156) | <2.2E-308 | Rare gastroenterologic disease | 8493 | 1059 | 2.533 (2.528 - 2.536) | <2.2E-308 |
| Rare hematologic disease | 37579 | 2269 | 3.05 (3.047 - 3.052) | <2.2E-308 | Rare bone diseases | 4113 | 476 | 2.351 (2.343 - 2.36) | <2.2E-308 |
| Rare infectious disease | 19326 | 1140 | 2.98 (2.975 - 2.985) | <2.2E-308 | Rare renal disease | 3226 | 368 | 2.317 (2.305 - 2.327) | <2.2E-308 |
| Rare systemic or rheumatologic disease | 47375 | 2526 | 2.693 (2.692 - 2.696) | <2.2E-308 | Rare ophthalmic disorder | 9767 | 1111 | 2.311 (2.308 - 2.315) | <2.2E-308 |
| Rare cardiac diseases | 3584 | 186 | 2.621 (2.596 - 2.649) | <2.2E-308 | Rare hepatic disease | 8375 | 944 | 2.29 (2.287 - 2.295) | <2.2E-308 |
| Rare maxillo facial surgical disease | 119 | <20 | 2.547 (1.866 - 3.476) | 1.88E-02 | Rare systemic or rheumatologic disease | 47375 | 5103 | 2.188 (2.187 - 2.189) | 1.88E-02 |
| Rare ophthalmic disorder | 9767 | 481 | 2.488 (2.477 - 2.497) | <2.2E-308 | Rare infectious disease | 19326 | 2077 | 2.183 (2.182 - 2.186) | <2.2E-308 |
| Rare immune disease | 9479 | 453 | 2.414 (2.403 - 2.423) | <2.2E-308 | Rare cardiac diseases | 3584 | 377 | 2.137 (2.126 - 2.146) | <2.2E-308 |
| Rare neurologic disease | 79021 | 3744 | 2.393 (2.393 - 2.395) | <2.2E-308 | Rare neurologic disease | 79021 | 8177 | 2.102 (2.102 - 2.103) | <2.2E-308 |
| Rare circulatory system disease | 727 | 33 | 2.293 (2.167 - 2.427) | 1.07E-06 | Rare developmental defect during embryogenesis | 25905 | 2641 | 2.071 (2.07 - 2.072) | <2.2E-308 |
| Rare renal disease | 3226 | 131 | 2.051 (2.021 - 2.08) | <2.2E-308 | Rare immune disease | 9479 | 868 | 1.86 (1.857 - 1.865) | <2.2E-308 |
| Rare developmental defect during embryogenesis | 25905 | 853 | 1.663 (1.66 - 1.667) | <2.2E-308 | Rare otorhinolaryngologic disease | 4875 | 432 | 1.8 (1.793 - 1.808) | <2.2E-308 |
| Rare inborn errors of metabolism | 6087 | 187 | 1.552 (1.537 - 1.569) | 1.08E-09 | Rare inborn errors of metabolism | 6087 | 492 | 1.642 (1.636 - 1.648) | <2.2E-308 |
| Rare otorhinolaryngologic disease | 4875 | 102 | 1.057 (1.037 - 1.077) | 5.73E-01 | Rare maxillo facial surgical disease | 119 | <20 | 1.366 (1.086 - 1.719) | 3.61E-01 |
| Rare surgical thoracis disease | 101 | <20 | 1.0 (0.383 - 2.614) | 1.00E+00 | Rare gynecologic or obstetric disease | 8419 | 515 | 1.243 (1.239 - 1.248) | 3.70E-07 |
| Rare urogenital disease | 9576 | 169 | 0.891 (0.881 - 0.902) | 0.1321632 | Rare urogenital disease | 9576 | 501 | 1.063 (1.059 - 1.067) | 1.61E-01 |
| Rare skin disease | 103614 | 1690 | 0.824 (0.823 - 0.825) | 1.78E-15 | Rare surgical thoracis disease | 101 | <20 | 1.006 (0.692 - 1.462) | 9.90E-01 |
| Rare gynecologic or obstetric disease | 8419 | 39 | 0.234 (0.223 - 0.246) | <2.2E-308 | Rare skin disease | 103614 | 4762 | 0.934 (0.934 - 0.935) | 1.62E-06 |

## Discussion

Although numerous research articles have been published on identifying rare disease patients in electronic health record (EHR) systems, there remains a notable scarcity of automated approaches capable of identifying a wide array of rare diseases on a large scale. To address this gap, we have developed an algorithm to automate the identification process of rare diseases in extensive EHR systems. The major outcome of this study is thus the development of a phenotyping algorithm that produces RD-specific codes, specifically 12,081 SNOMED-CT codes and 357 ICD-10 codes representing at least 6,342 RDs. To the best of our knowledge, this is the largest number of RDs represented by SNOMED-CT and ICD-10 codes. These RDs represent 30 Orphanet classes (**Supplementary Table 4**). The availability of these codes allows investigation of RDs as groups, rather than individuals or small numbers of RDs, in any EHR system. This capacity empowers clinicians and researchers to find novel shared treatment options for this difficult to study population.

Our phenotype algorithm addresses major challenges in systematically defining RDs in EHR systems. First, we eliminated RD codes that relate to group of disorders that could erroneously bring in common diseases when we expanded the code list with their descendants, and we also removed those that relate to phenotypes. Second, we focused on minimizing manual curation so that the process can be readily executed when updates to source databases (e.g., GARD, Orphanet, OHDSI) are applicable. Third, recognizing the lack of convergence in the definition of rare diseases across the world, we anchored our algorithm to GARD IDs and thus followed the US definition of RDs. While our algorithm produces a comprehensive list of RD codes, we acknowledge that some RDs might be missed because of the exclusion criteria applied. In addition, as we minimize manual curation, we recommend careful evaluation of codes when they are used to create a cohort. In our study, we added an incidence filter to remove potentially erroneously represented RDs in the EHR system. This filter removed 25 codes (**Supplementary Table 5**) which represent common diseases. The remaining are likely to either represent issues in how the codes are used in the EHR systems or could represent false positives (a disease we consider rare but is actually common).

A recent study by Thygesen et al^17^ evaluated the prevalence, and clinical and demographic data for RD using a large retrospective observational study in England. An algorithm specific to their study was designed to identify RD using Orphanet and implement filters to remove certain disease types (e.g., clinical group, clinical subtype, etiological subtype, and biological anomaly, etc.) and mappings without existing codes in the observational study. While some key similarities exist between the algorithm presented in Thygesen et al. and ours (e.g., use of Orphanet, incidence and removal or groups/broad disorders), our algorithm produces codes that are independent of a specific cohort. Therefore, our list of RD-specific codes can be used directly as a starting point for any EHR system.

We note that there is a large skew in the distribution of the numbers of RD-specific codes and associated publications by linearization classes (**Figure 3**). Surprisingly, while 80% of RDs are related to genetic disorders, only 3 RD-specific codes map to that category^33^. Notably, this underrepresentation is an artifact of the rare genetic disease linearization and the priority rules applied when mapping a disease to a linearization^30^. Specifically for rare genetic diseases, a disease is first mapped to the most relevant body system affected, with involvement for prognosis and specialist type for disease management. The remaining diseases only will be mapped to rare genetic disease. If this linearization were to be further analyzed, the grouping of this genetic RD category could be revised.

As a use case of this list of RD-specific codes, we systematically identified RD patients in a COVID-19 cohort established within the N3C enclave to broadly compare the demographics of COVID-19 patients with and without preexisting RDs and to evaluate the RRs of COVID-19 related hospitalization and mortality in patients with and without preexisting RDs. Our analyses are preliminary, as only univariate analyses were performed, and we did not adjust for relevant clinical covariables, which is beyond the scope of this manuscript. Nonetheless, this analysis exemplifies the utility of our RD-specific codes and represents, to the best of our knowledge, the largest study in the United States that evaluates COVID-19 severity outcomes in relation to RD classes at scale by maximizing the coverage of RD patients using a comprehensive list of RD-related codes. We provided unadjusted risk ratios of each rare disease class with respect to COVID-19 positive patients without rare diseases. Similar to our findings, Thygesen et al. reported that patients with rare developmental defect during embryogenesis, rare renal disease, rare neurologic disease, rare immune disease, rare ophthalmic disorder, rare systemic or rheumatologic disease, and rare bone diseases had higher risk of mortality in their cohort of COVID-19 patients that were vaccinated. They also found that patients with rare genetic, rare and systemic and rheumatological diseases of childhood showed a higher risk of mortality although those RD groups were not evaluated in our study as they were not represented in our cohort. Thygesen et al. also reported that patients with rare skin disease showed a higher risk of mortality while in our analyses, this group exhibited a protective effect. This discrepancy could be due to differences in the diseases represented in both cohorts.

Our results also align with a comprehensive analysis in Hong Kong showing that RD patients had an increased risk of COVID-19 related mortality compared with the general population^34^. Interestingly, stratified analyses in the Hong Kong cohort demonstrated that the risk of mortality increases with age in the general population whereas in the RD population, mortality rates are much higher in the younger population (≤18 years old). In our study, we did not find a large increase in the percent mortality for RD patients in the younger population (2-20 years) although we did find a higher percent mortality in the RD population compared to the non-RD population. We note here that our study does not account for vaccination status, exposure to anti-virals, demographics and other relevant clinical factors since the focus of the study is on the generation of RD-specific codes. Findings from our preliminary use case demonstrate the utility of our RD-specific codes in identifying RD patients in large clinical cohorts. As we know, RD patients are underrepresented in EHR systems due to heterogeneity of the coding systems used in different clinical settings. Our approach is based on Orphanet classification and further usage of ICD-10 codes or SNOMED-CT codes to the confirmed cases of rare diseases. Patients with suspected rare diseases or undiagnosed conditions were not included in our analysis. Further limitation, The N3C (National COVID Cohort Collaborative) data are aggregated from multiple healthcare systems, which use four common data models with varying levels of granularity. However, harmonizing these disparate data requires making assumptions and inferences, which could introduce systematic biases. Additionally, accurately determining race within N3C is challenging due to variations in how race is reported across different healthcare systems.

We also recognize that ICD-10 codes lack granularity in RDs. Orphanet’s mappings currently cover only three-digit ICD-10 codes rather than the ICD10-CM four-digit level codes that provide more precision. Once these become available, our algorithm could be readily adapted to use ICD10-CM rather than ICD-10. The mappings could be improved even further by leveraging ICD-11 codes alongside SNOMED-CT.

Overall, these findings provide a robust foundation for broader evaluation of RDs in EHR systems. Evaluations of groups of RDs are advantageous for increasing the statistical power for finding clinically relevant patterns and better understanding RDs as complex systems rather than single diseases. Our use case evaluation in COVID-19 provided preliminary findings that further highlight the need for tailored monitoring of RD patients to prevent worse COVID-19 outcomes. Moving forward, this framework provides a crucial foundation for future investigations into how comorbidities, vaccination status, and antiviral treatments influence outcomes such as COVID-19-related mortality, hospitalization, long COVID, and reinfection among individuals with preexisting rare diseases. Continued research in this direction holds significant potential to improve clinical decision-making and health equity for this often-overlooked population.

## Data Availability

The analyses described in this [abstract/publication/report/presentation] were conducted with data or tools accessed through the NCATS N3C Data Enclave https://covid.cd2h.org and N3C Attribution & Publication Policy v 1.2-2020-08-25b supported by NCATS Contract No. 75N95023D00001, Axle Informatics Subcontract: NCATS-P00438-B, and [insert additional funding agencies or sources and reference numbers as declared by the contributors in their form response above]. This research was possible because of the patients whose information is included within the data and the organizations (https://ncats.nih.gov/n3c/resources/data-contribution/data-transfer-agreement-signatories) and scientists who have contributed to the on-going development of this community resource [https://doi.org/10.1093/jamia/ocaa196].

## Funding

This work was supported in part by the intramural and extramural programs at NCATS (ZIA ZICTR000410).

## Authors Contributions

EAM and ASY conceived and designed the study. ASY performed all data analyses. ASY and HS collaboratively validated concept descendants for selected diseases. ASY and EAM interpreted the results and drafted the manuscript. QZ and ES provided substantial revisions and editorial input. QZ, ES, CZ, ASY, and EAM curated disease lists. All authors reviewed and approved the final manuscript and agreed to be accountable for all aspects of the work.

See https://github.com/arjunyadaw/Rare-Disease-Cohort-Building-in-EHR-System.git

## Competing Interests

The authors declare that they have no competing interests.

## National COVID Cohort Collaborative attribution

### Disclaimer

The N3C Publication committee confirmed that this [manuscript/abstract/poster/presentation] MSID:2402.984 is in accordance with N3C data use and attribution policies; however, this content is solely the responsibility of the authors and does not necessarily represent the official views of the National Institutes of Health or the N3C program.

## Institutional review board

The N3C data transfer to NCATS is performed under a Johns Hopkins University Reliance Protocol # IRB00249128 or individual site agreements with NIH. The N3C Data Enclave is managed under the authority of the NIH; information can be found at https://ncats.nih.gov/n3c/resources.

## Individual acknowledgments for core contributors

We gratefully acknowledge the following core contributors to N3C: Adam B. Wilcox, Adam M. Lee, Alexis Graves, Alfred (Jerrod) Anzalone, Amin Manna, Amit Saha, Amy Olex, Andrea Zhou, Andrew E. Williams, Andrew M. Southerland, Andrew T. Girvin, Anita Walden, Anjali Sharathkumar, Benjamin Amor, Benjamin Bates, Brian Hendricks, Brijesh Patel, G. Caleb Alexander, Carolyn T. Bramante, Cavin Ward-Caviness, Charisse Madlock-Brown, Christine Suver, Christopher G. Chute, Christopher Dillon, Chunlei Wu, Clare Schmitt, Cliff Takemoto, Dan Housman, Davera Gabriel, David A. Eichmann, Diego Mazzotti, Donald E. Brown, Eilis Boudreau, Elaine L. Hill, Emily Carlson Marti, Emily R. Pfaff, Evan French, Farrukh M Koraishy, Federico Mariona, Fred Prior, George Sokos, Greg Martin, Harold P. Lehmann, Heidi Spratt, Hemalkumar B. Mehta, J.W. Awori Hayanga, Jami Pincavitch, Jaylyn Clark, Jeremy Richard Harper, Jessica Yasmine Islam, Jin Ge, Joel Gagnier, Johanna J. Loomba, John B. Buse, Jomol Mathew, Joni L. Rutter, Julie A. McMurry, Justin Guinney, Justin Starren, Karen Crowley, Katie Rebecca Bradwell, Kellie M. Walters, Ken Wilkins, Kenneth R. Gersing, Kenrick Cato, Kimberly Murray, Kristin Kostka, Lavance Northington, Lee Pyles, Lesley Cottrell, Lili M. Portilla, Mariam Deacy, Mark M. Bissell, Marshall Clark, Mary Emmett, Matvey B. Palchuk, Melissa A. Haendel, Meredith Adams, Meredith Temple-O’Connor, Michael G. Kurilla, Michele Morris, Nasia Safdar, Nicole Garbarini, Noha Sharafeldin, Ofer Sadan, Patricia A. Francis, Penny Wung Burgoon, Philip R.O. Payne, Randeep Jawa, Rebecca Erwin-Cohen, Rena C. Patel, Richard A. Moffitt, Richard L. Zhu, Rishikesan Kamaleswaran, Robert Hurley, Robert T. Miller, Saiju Pyarajan, Sam G. Michael, Samuel Bozzette, Sandeep K. Mallipattu, Satyanarayana Vedula, Scott Chapman, Shawn T. O’Neil, Soko Setoguchi, Stephanie S. Hong, Steven G. Johnson, Tellen D. Bennett, Tiffany J. Callahan, Umit Topaloglu, Valery Gordon, Vignesh Subbian, Warren A. Kibbe, Wenndy Hernandez, Will Beasley, Will Cooper, William Hillegass, Xiaohan Tanner Zhang. Details of contributions available at covid.cd2h.org/core-contributors Note: Google Scholar’s indexing accuracy can be compromised due to factors like language mismatches in text and metadata, incorrect author name formatting, and errors in automated bibliographic data extraction. Indexing may also take 6–8 weeks post-publication.

## Data Partners with Released Data

The following institutions whose data is released or pending:

Available: Advocate Health Care Network — UL1TR002389: The Institute for Translational Medicine (ITM) • Aurora Health Care Inc — UL1TR002373: Wisconsin Network For Health Research • Boston University Medical Campus — UL1TR001430: Boston University Clinical and Translational Science Institute • Brown University — U54GM115677: Advance Clinical Translational Research (Advance-CTR) • Carilion Clinic — UL1TR003015: iTHRIV Integrated Translational health Research Institute of Virginia • Case Western Reserve University — UL1TR002548: The Clinical & Translational Science Collaborative of Cleveland (CTSC) • Charleston Area Medical Center — U54GM104942: West Virginia Clinical and Translational Science Institute (WVCTSI) • Children’s Hospital Colorado — UL1TR002535: Colorado Clinical and Translational Sciences Institute • Columbia University Irving Medical Center — UL1TR001873: Irving Institute for Clinical and Translational Research • Dartmouth College — None (Voluntary) Duke University — UL1TR002553: Duke Clinical and Translational Science Institute • George Washington Children’s Research Institute — UL1TR001876: Clinical and Translational Science Institute at Children’s National (CTSA-CN) • George Washington University — UL1TR001876: Clinical and Translational Science Institute at Children’s National (CTSA-CN) • Harvard Medical School — UL1TR002541: Harvard Catalyst • Indiana University School of Medicine — UL1TR002529: Indiana Clinical and Translational Science Institute • Johns Hopkins University — UL1TR003098: Johns Hopkins Institute for Clinical and Translational Research • Louisiana Public Health Institute — None (Voluntary) • Loyola Medicine — Loyola University Medical Center • Loyola University Medical Center — UL1TR002389: The Institute for Translational Medicine (ITM) • Maine Medical Center — U54GM115516: Northern New England Clinical & Translational Research (NNE-CTR) Network • Mary Hitchcock Memorial Hospital & Dartmouth Hitchcock Clinic — None (Voluntary) • Massachusetts General Brigham — UL1TR002541: Harvard Catalyst • Mayo Clinic Rochester — UL1TR002377: Mayo Clinic Center for Clinical and Translational Science (CCaTS) • Medical University of South Carolina — UL1TR001450: South Carolina Clinical & Translational Research Institute (SCTR) • MITRE Corporation — None (Voluntary) • Montefiore Medical Center — UL1TR002556: Institute for Clinical and Translational Research at Einstein and Montefiore • Nemours — U54GM104941: Delaware CTR ACCEL Program • NorthShore University HealthSystem — UL1TR002389: The Institute for Translational Medicine (ITM) • Northwestern University at Chicago — UL1TR001422: Northwestern University Clinical and Translational Science Institute (NUCATS) • OCHIN — INV-018455: Bill and Melinda Gates Foundation grant to Sage Bionetworks • Oregon Health & Science University — UL1TR002369: Oregon Clinical and Translational Research Institute • Penn State Health Milton S. Hershey Medical Center — UL1TR002014: Penn State Clinical and Translational Science Institute • Rush University Medical Center — UL1TR002389: The Institute for Translational Medicine (ITM) • Rutgers, The State University of New Jersey — UL1TR003017: New Jersey Alliance for Clinical and Translational Science • Stony Brook University — U24TR002306 • The Alliance at the University of Puerto Rico, Medical Sciences Campus — U54GM133807: Hispanic Alliance for Clinical and Translational Research (The Alliance) • The Ohio State University — UL1TR002733: Center for Clinical and Translational Science • The State University of New York at Buffalo — UL1TR001412: Clinical and Translational Science Institute • The University of Chicago — UL1TR002389: The Institute for Translational Medicine (ITM) • The University of Iowa — UL1TR002537: Institute for Clinical and Translational Science • The University of Miami Leonard M. Miller School of Medicine — UL1TR002736: University of Miami Clinical and Translational Science Institute • The University of Michigan at Ann Arbor — UL1TR002240: Michigan Institute for Clinical and Health Research • The University of Texas Health Science Center at Houston — UL1TR003167: Center for Clinical and Translational Sciences (CCTS) • The University of Texas Medical Branch at Galveston — UL1TR001439: The Institute for Translational Sciences • The University of Utah — UL1TR002538: Uhealth Center for Clinical and Translational Science • Tufts Medical Center — UL1TR002544: Tufts Clinical and Translational Science Institute • Tulane University — UL1TR003096: Center for Clinical and Translational Science • The Queens Medical Center — None (Voluntary) • University Medical Center New Orleans — U54GM104940: Louisiana Clinical and Translational Science (LA CaTS) Center • University of Alabama at Birmingham — UL1TR003096: Center for Clinical and Translational Science • University of Arkansas for Medical Sciences — UL1TR003107: UAMS Translational Research Institute • University of Cincinnati — UL1TR001425: Center for Clinical and Translational Science and Training • University of Colorado Denver, Anschutz Medical Campus — UL1TR002535: Colorado Clinical and Translational Sciences Institute • University of Illinois at Chicago — UL1TR002003: UIC Center for Clinical and Translational Science • University of Kansas Medical Center — UL1TR002366: Frontiers: University of Kansas Clinical and Translational Science Institute • University of Kentucky — UL1TR001998: UK Center for Clinical and Translational Science • University of Massachusetts Medical School Worcester — UL1TR001453: The UMass Center for Clinical and Translational Science (UMCCTS) • University Medical Center of Southern Nevada — None (voluntary) • University of Minnesota — UL1TR002494: Clinical and Translational Science Institute • University of Mississippi Medical Center — U54GM115428: Mississippi Center for Clinical and Translational Research (CCTR) • University of Nebraska Medical Center — U54GM115458: Great Plains IDeA-Clinical & Translational Research • University of North Carolina at Chapel Hill — UL1TR002489: North Carolina Translational and Clinical Science Institute • University of Oklahoma Health Sciences Center — U54GM104938: Oklahoma Clinical and Translational Science Institute (OCTSI) • University of Pittsburgh — UL1TR001857: The Clinical and Translational Science Institute (CTSI) • University of Pennsylvania — UL1TR001878: Institute for Translational Medicine and Therapeutics • University of Rochester — UL1TR002001: UR Clinical & Translational Science Institute • University of Southern California — UL1TR001855: The Southern California Clinical and Translational Science Institute (SC CTSI) • University of Vermont — U54GM115516: Northern New England Clinical & Translational Research (NNE-CTR) Network • University of Virginia — UL1TR003015: iTHRIV Integrated Translational health Research Institute of Virginia • University of Washington — UL1TR002319: Institute of Translational Health Sciences • University of Wisconsin-Madison — UL1TR002373: UW Institute for Clinical and Translational Research • Vanderbilt University Medical Center — UL1TR002243: Vanderbilt Institute for Clinical and Translational Research • Virginia Commonwealth University — UL1TR002649: C. Kenneth and Dianne Wright Center for Clinical and Translational Research • Wake Forest University Health Sciences — UL1TR001420: Wake Forest Clinical and Translational Science Institute • Washington University in St. Louis — UL1TR002345: Institute of Clinical and Translational Sciences • Weill Medical College of Cornell University — UL1TR002384: Weill Cornell Medicine Clinical and Translational Science Center • West Virginia University — U54GM104942: West Virginia Clinical and Translational Science Institute (WVCTSI) Submitted: Icahn School of Medicine at Mount Sinai — UL1TR001433: ConduITS Institute for Translational Sciences • The University of Texas Health Science Center at Tyler — UL1TR003167: Center for Clinical and Translational Sciences (CCTS) • University of California, Davis — UL1TR001860: UCDavis Health Clinical and Translational Science Center • University of California, Irvine — UL1TR001414: The UC Irvine Institute for Clinical and Translational Science (ICTS) • University of California, Los Angeles — UL1TR001881: UCLA Clinical Translational Science Institute • University of California, San Diego — UL1TR001442: Altman Clinical and Translational Research Institute • University of California, San Francisco — UL1TR001872: UCSF Clinical and Translational Science Institute NYU Langone Health Clinical Science Core, Data Resource Core, and PASC Biorepository Core — OTA-21-015A: Post-Acute Sequelae of SARS-CoV-2 Infection Initiative (RECOVER) Pending: Arkansas Children’s Hospital — UL1TR003107: UAMS Translational Research Institute • Baylor College of Medicine — None (Voluntary) • Children’s Hospital of Philadelphia — UL1TR001878: Institute for Translational Medicine and Therapeutics • Cincinnati Children’s Hospital Medical Center — UL1TR001425: Center for Clinical and Translational Science and Training • Emory University — UL1TR002378: Georgia Clinical and Translational Science Alliance • HonorHealth — None (Voluntary) • Loyola University Chicago — UL1TR002389: The Institute for Translational Medicine (ITM) • Medical College of Wisconsin — UL1TR001436: Clinical and Translational Science Institute of Southeast Wisconsin • MedStar Health Research Institute — None (Voluntary) • Georgetown University — UL1TR001409: The Georgetown-Howard Universities Center for Clinical and Translational Science (GHUCCTS) • MetroHealth — None (Voluntary) • Montana State University — U54GM115371: American Indian/Alaska Native CTR • NYU Langone Medical Center — UL1TR001445: Langone Health’s Clinical and Translational Science Institute • Ochsner Medical Center — U54GM104940: Louisiana Clinical and Translational Science (LA CaTS) Center • Regenstrief Institute — UL1TR002529: Indiana Clinical and Translational Science Institute • Sanford Research — None (Voluntary) • Stanford University — UL1TR003142: Spectrum: The Stanford Center for Clinical and Translational Research and Education • The Rockefeller University — UL1TR001866: Center for Clinical and Translational Science • The Scripps Research Institute — UL1TR002550: Scripps Research Translational Institute • University of Florida — UL1TR001427: UF Clinical and Translational Science Institute • University of New Mexico Health Sciences Center — UL1TR001449: University of New Mexico Clinical and Translational Science Center • University of Texas Health Science Center at San Antonio — UL1TR002645: Institute for Integration of Medicine and Science • Yale New Haven Hospital — UL1TR001863: Yale Center for Clinical Investigation

## Notes

### Competing Interest Statement

The authors have declared no competing interest.

### Author Declarations

In this study, We have used N3C enclave data which is accessible by researcher by signing DUA (https://covid.cd2h.org). The N3C Data Enclave is approved under the authority of the National Institutes of Health Institutional Review Board. Each N3C site maintains an institutional review board–approved data transfer agreement.

