## Supplementary File for "Systematic identification of rare disease patients in electronic health records enables evaluation of clinical outcomes"

**Method (mapping rare disease patients in EHR systems):**

We constructed a cohort of COVID-19 infected individuals that had a rare disease (RDs) diagnosis prior to COVID-19 infection (**Supplementary Figure 2**). To accomplish this, we created a curated list of unique rare diseases, defined by 12,003 GARD IDs. These GARD IDs were mapped with ORPHANET (July 2023 version) which mapped 9,369 rare diseases with GARD IDs and 2,634 GARD IDs excluded due to no mapping to ORPHANET. Of the 9,369 GARD IDs mapping to ORPHANET, were represented by 6,553 SNOMED-CT and 564 ICD-10 codes. We note that ICD-10 codes represent single diseases with no descendants.

The remaining GARD IDs were subject to a series of exclusion criteria and filters:

1) Groups of disorder exclusion: 6,553 rare diseases mapped from ORPHANET to SNOEMD-CT and 575 rare diseases mapped from ORPHANET to ICD-10 codes. There is 226 SNOMED-CT RDs excluded and 101 ICD-10 RDs excluded due to the group of disorders which results in 6,327 SNOMED-CT codes and 463 ICD-10 codes.
2) SNOMED-CT mapping to OHDSI: 6,327 RDs with SNOMED-CT codes mapped to OHDSI atlas which brought a total of 12,569 descendent concept ids along with parent disease node with vocabulary: SNOMED and domain: Condition criteria.
3) Phenotype Filter: are associated with rare disease with descendent concepts phenotype mapping for SNOMED-CT and ICD-10 codes mapping with HPO. There are 106 ICD-10 codes, and 488 SNOMED-CT codes excluded due to phenotype mapping. Now remaining diseases with ICD-10/SNOMED-CT codes mapped at N3C enclave. Finally, 12,081 SNOMED-CT codes and 357 ICD-10 codes for further classification **(Figure2)**.
4) Orphanet Linearization: To avoid the principle of polyhierarchy, rare diseases in the Orphanet database are sorted by medical specialty, ensuring that each disease belongs to only one specialty. We created rare disease classes based on ORPHANET linearization which classified rare diseases into 30 rare disease classes (**Supplementary Table 4**). Each rare disease class follows the same mapping flow which is described in flow diagram figure (**Figure2**) to get final concept IDs based on ICD-10 code and/or SNOMED-CT codes. Each rare disease with GARD IDs and ORPHACode mapped with SNOMED-CT /ICD-10 codes are linearized into 30 classes (listed in (**Supplementary Table 2 & 3**). Rare genetic disease (ORPHA98053) must not be used as a linearization parent. When a disease is included in multiple classifications corresponding to various medical specialties, priority is given to the specialty:

- corresponding to the most severely affected body system.
- corresponding with the most determining involvement for the prognosis.
- corresponding with the specialist most likely to be relied on for the management of the disease.

5) Rare surgical cardiac disease and rare genetic disease class dropped due to no ICD-10 mapping and either no SNOMED-CT code or SNOMED-CT code mapped to phenotype filtering. Also, rare infertility diseases have no patient mapping at N3C enclave.
6) Four rare disease classes 1) “rare disease due to toxic affect” removed because concepts under this class were associated with uncommon toxic exposures (e.g., Cyanide, Snake Venom etc.). 2) rare transplantation disease, are directly a comorbidity or complication of having had transplantation etc. 3) “rare odontologic disease” (37 patients) with no mortality; 4) “rare abdominal surgical diseases” (4 patients) with no mortality. So, a total of seven rare disease classes were not considered for further study out of 30 classes.
7) Finally, there are 23 rare disease classes considered for further study of COVID-19 positive cohorts with preexisting rare diseases.
8) Incidence Filter: Is the incidence of each disease (with ICD-10 and/or SNOMED-CT) > 6/10,000? If not, then corresponding ICD-10 and SNOMED-CTs are retained. If yes, then diseases are excluded. There are 7 & 18 diseases dropped due to common/categorical from ICD-10/SNOMED-CT mapping and rest retained in the rare disease cohort (**Supplementary Figure 4**). Finally, 350 rare diseases with ICD-10 codes (**Supplementary Table 3**) and 12,063 rare diseases (or its descendants) with SNOMED-CT codes will be mapped to the OMOP concept table at N3C enclave (**Supplementary Table 2**).

After application of these filters and exclusion criteria, we created different classes of rare disease cohorts by following the same filtering and exclusion criteria which brought a total of 404,735 rare disease patients for 23 rare disease classes.

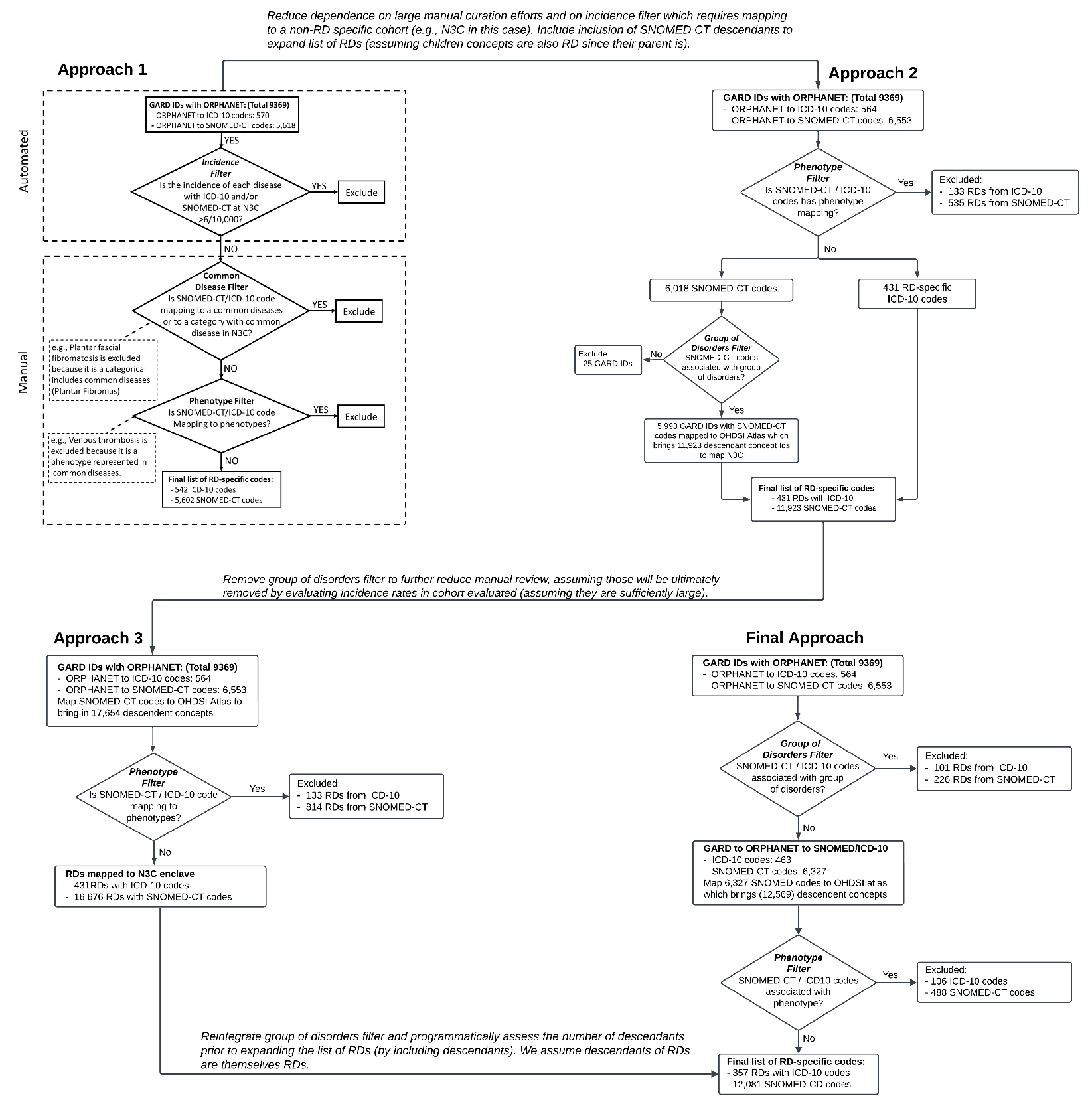

**Supplementary Figure 1: Evolution of semi-automated approaches of rare disease extraction from EHR systems by using ICD-10 and SMOCED-CT codes.** This approach started with 12,003 rare disease GARD IDs and followed approaches 1,2,3 and final approach to find SNOMED-CT and ICD-10 codes which minimized group of disorders and phenotype diseases.

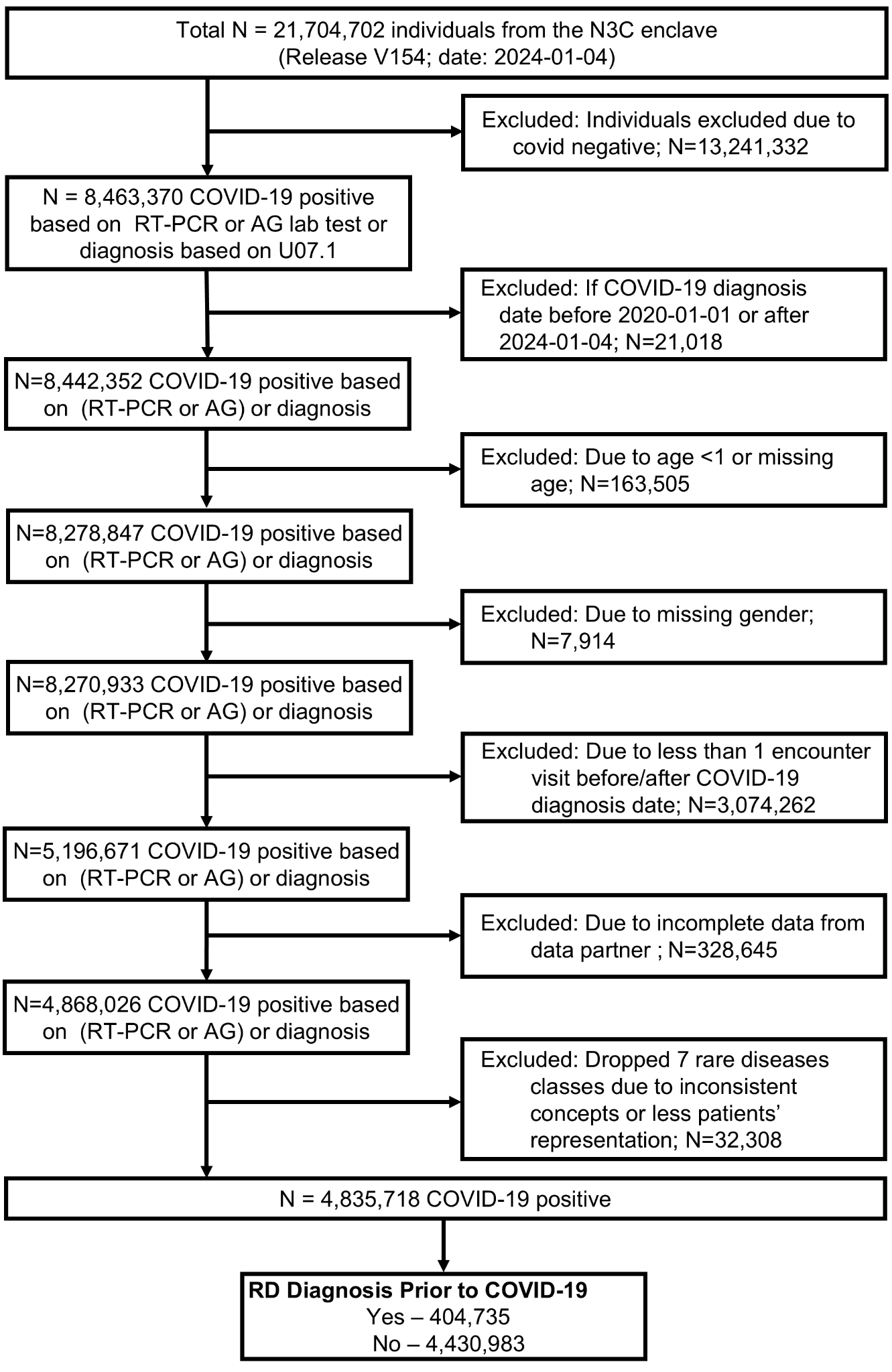

**Supplementary Figure 2: Workflow to define COVID-19 patients with and without RD in N3C.** Of the 21,704,702 patients in the N3C enclave release version V154, 4,835,718 were defined as COVID-19 positive between 2020-01-01 to 2024-01-04, as confirmed by a RT-PCR or Antigen test or diagnosis test. This subset also passed the following filters: non missing values for age and sex, at least one encounter visits before or after covid diagnosis date, incomplete data from data partners. See supplementary methods on detailed inclusion or exclusion criteria for cohort building. Lastly, patients with COVID-19 were stratified into two groups, those with and without RDs, based on our generated SNOMED-CT and ICD-10 codes.

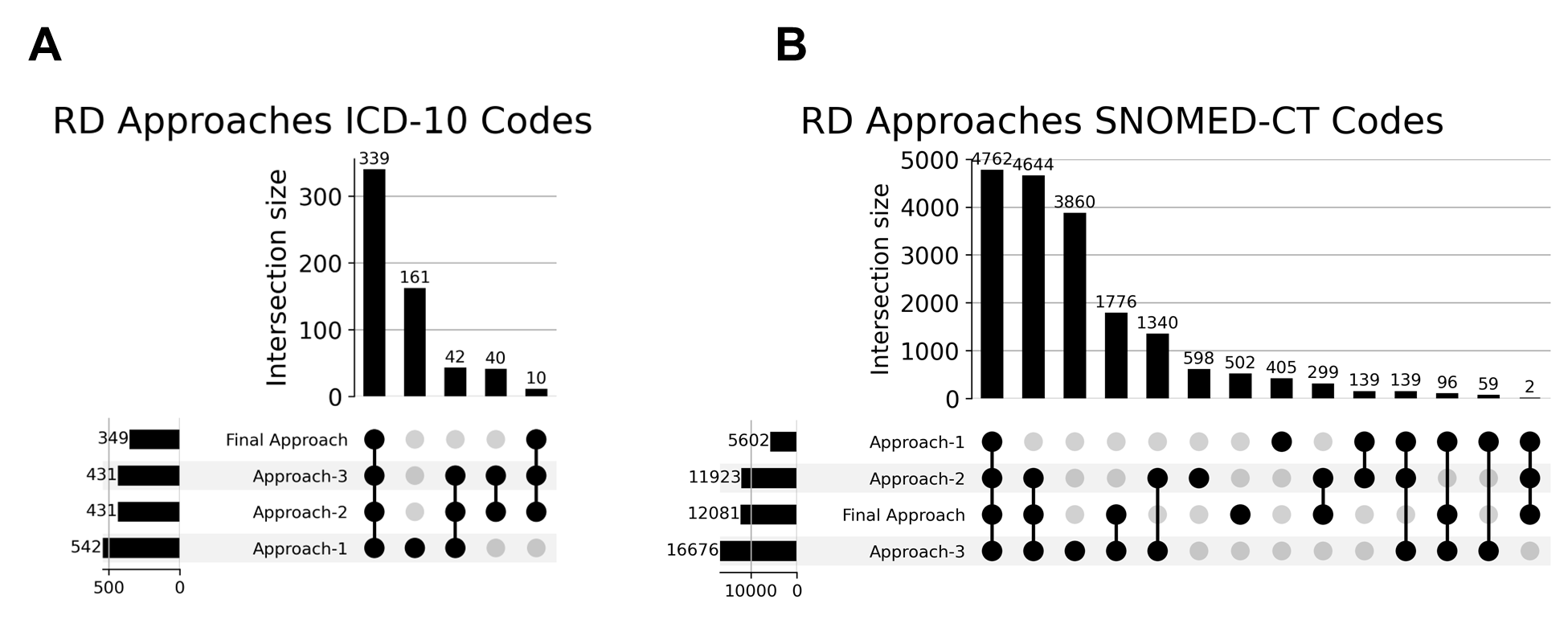

**Supplementary Figure 3: Upset plots of different approaches (ICD-10 and SNOMED-CT codes).** A) Upset plot shows number of ICD-10 codes common in all approaches where approach-1,2 & 3 had few redundant codes which were dropped due to group of disorders. B) Similarly for SNOMED-CD code more than 4500 SNOMED-CT codes are common amongst approaches-1,2,3 & final approach and many redundant codes were existing in approaches-1,2 & 3 due to groups of disorders or due to phenotype.

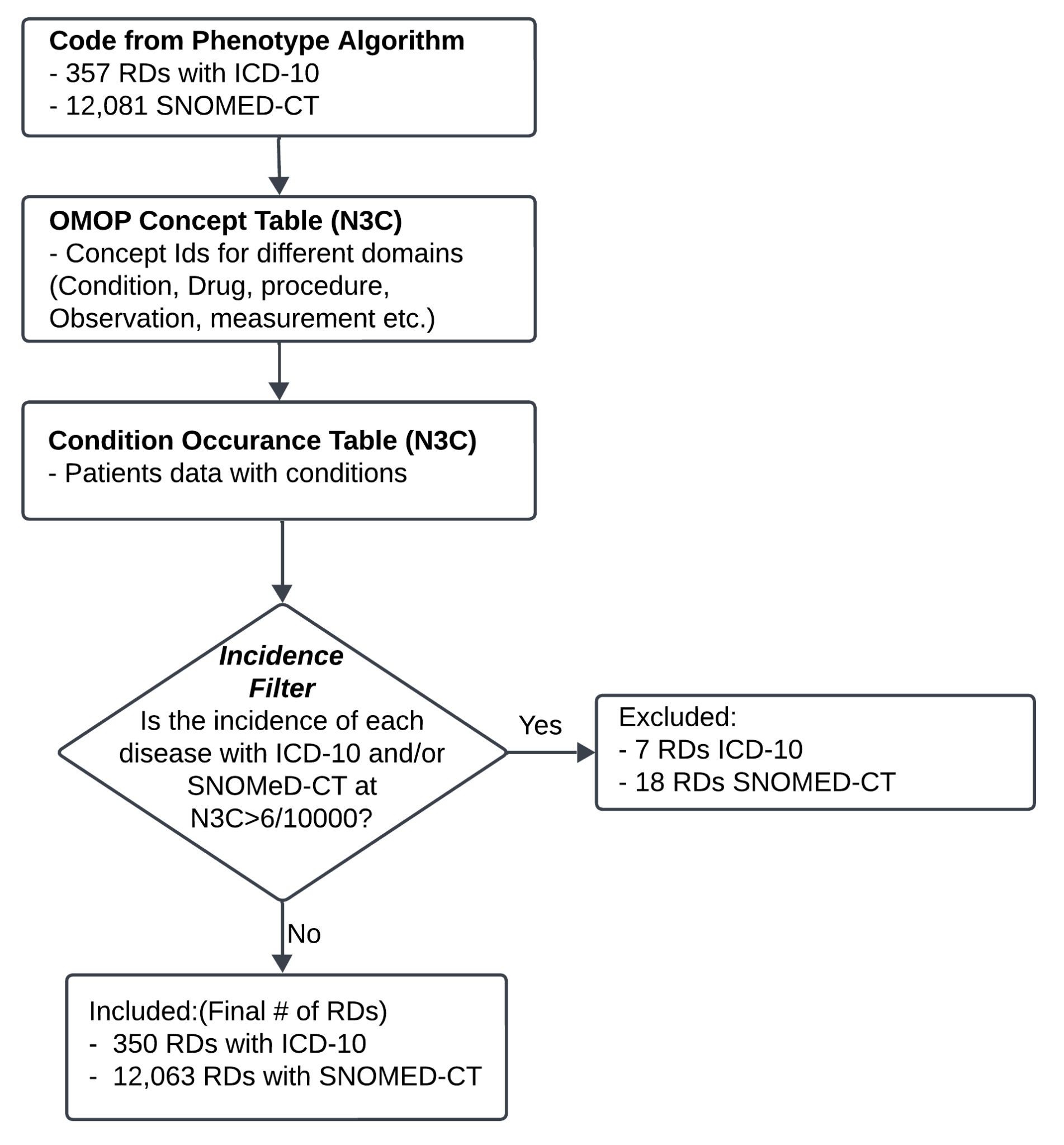

**Supplementary Figure 4:** **Incidence filter based on RD phenotyping Algorithm codes at N3C enclave.** A semi-automated algorithm was built to define RD-specific ICD-10 and SNOMED-CT codes. As a result, 350 ICD-10 codes and 12,063 SNOMED-CT codes are produced by phenotyping algorithm. These codes were then used to identify RD specific concept ids at OMOP concept table and then mapped to condition occurrence table at N3C enclave which produced RDs with associated patients. We used an additional filter based on the incidence of RD patients in N3C to exclude diseases that had a higher-than-expected incidence in the enclave.

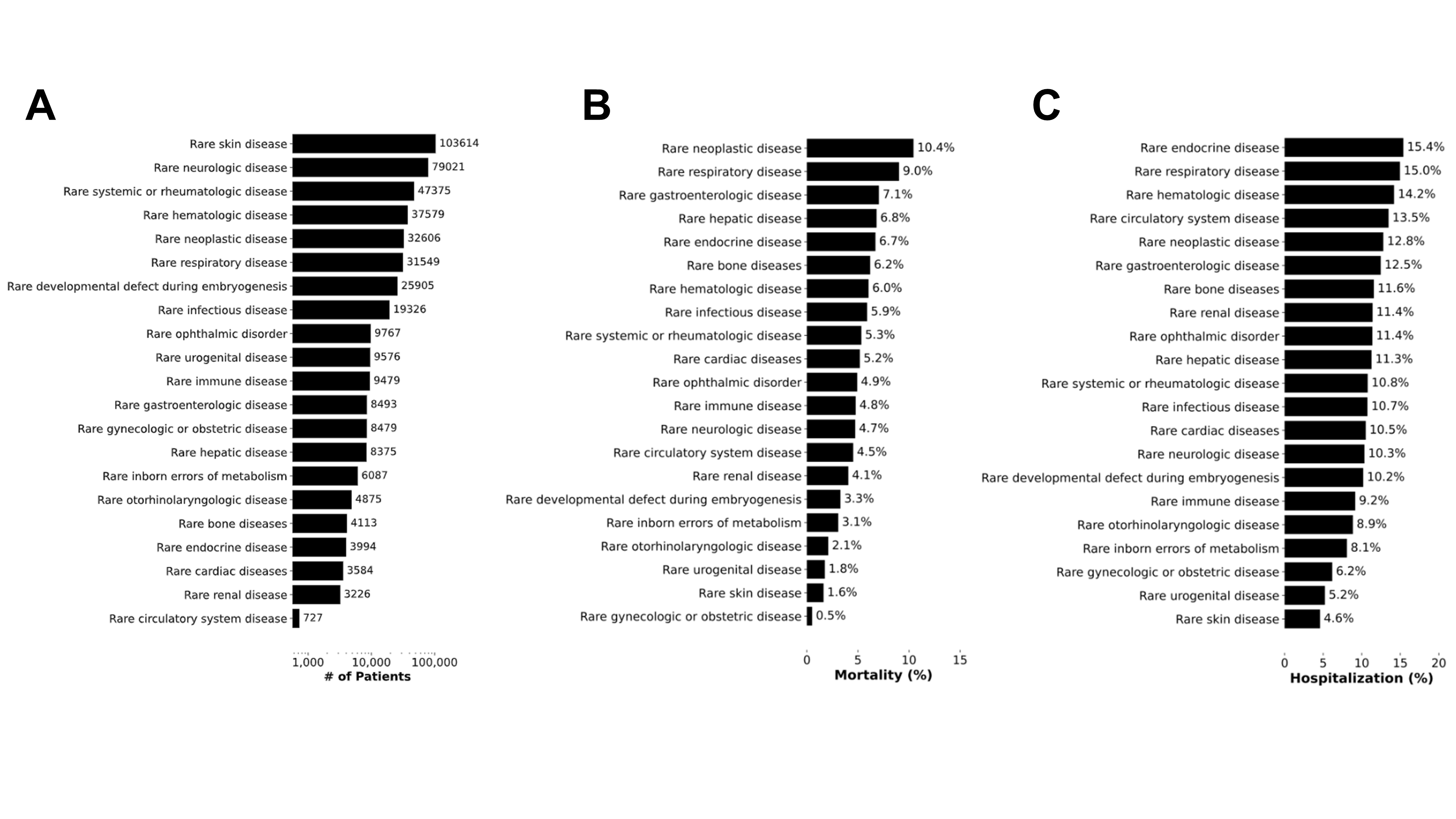

**Supplementary Figure 5: Description of patients with preexisting RD identified in the N3C COVID-19 cohort.** A) Number of patients per RD class for RD patients identified within our N3C COVID-19 cohort. B and C): Distribution of the number of RD patients with a COVID-19 diagnosis stratified by COVID-19 related mortality and hospitalization, respectively.

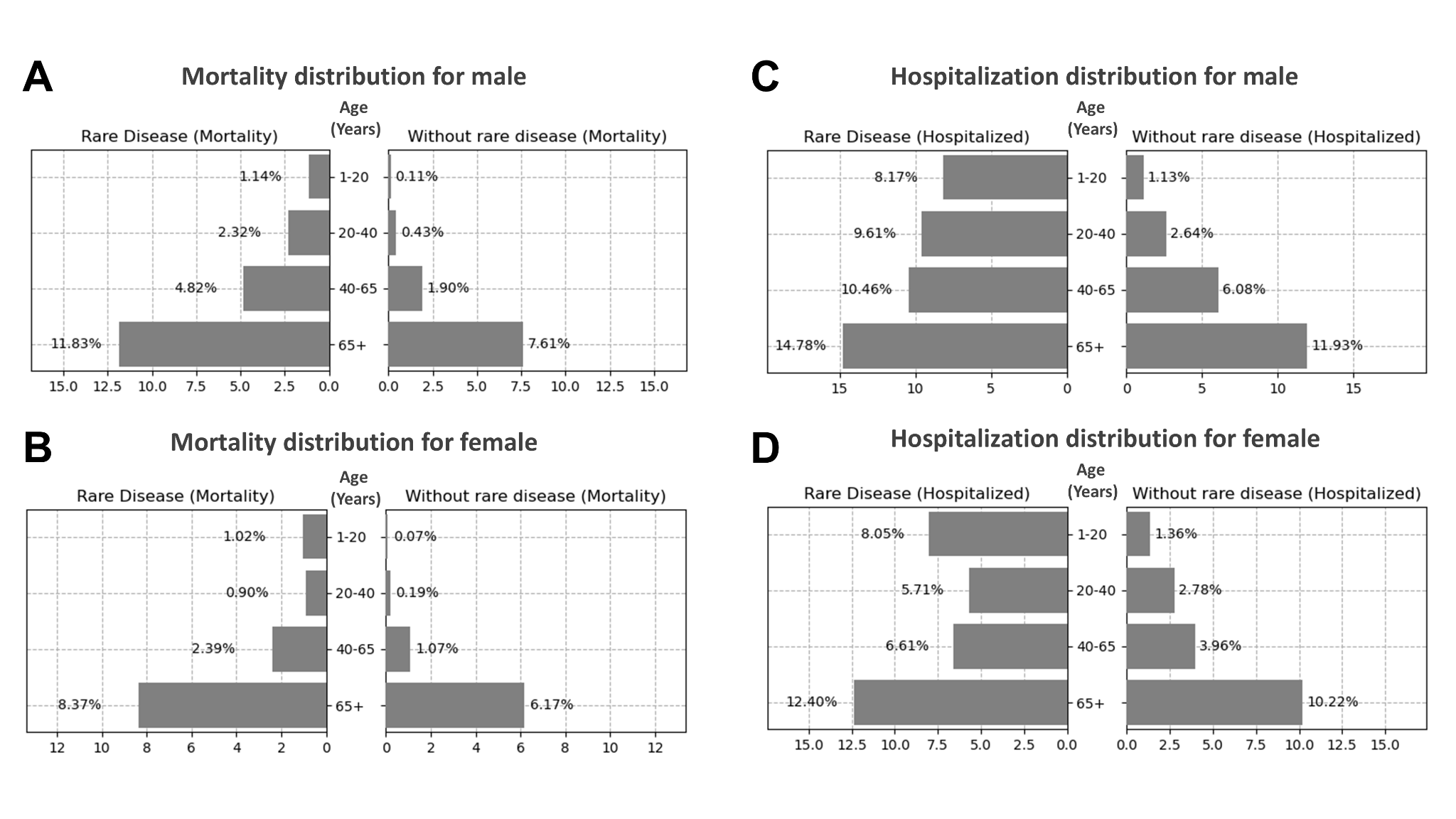

**Supplementary Figure 6: Distribution of the number of patients with preexisting RD that have suffered COVID-19 related death or hospitalization, stratified by age and sex.** A, B) Breakdown of COVID-19 patients' mortality with/without preexisting RD by sex male and female, respectively. (C, D) Breakdown of COVID-19 patients' hospitalization with/without preexisting RD by sex male and female, respectively.
